# High-Throughput Screening for Prescribing Cascades Among Real-World Angiotensin-II Receptor Blockers (ARBs) Initiators

**DOI:** 10.1101/2025.03.10.25323711

**Authors:** Asinamai M. Ndai, Kayla Smith, Shailina Keshwani, Jaeyoung Choi, Michael Luvera, Tanner Beachy, Marianna Calvet, Carl J. Pepine, Stephan Schmidt, Scott M. Vouri, Earl J. Morris, Steven M Smith

**Author notes:** Corresponding Author: Steven M Smith, PharmD, MPH, Department of Pharmaceutical Outcomes & Policy University of Florida, College of Pharmacy, PO Box 100496, Gainesville, FL 32611. **Prior Presentation:** Preliminary results of this study are presented in the American Heart Association Hypertension Scientific Session, 2023.

## Abstract

**Objective:** Angiotensin-II Receptor Blockers (ARBs) are commonly prescribed; however, their adverse events may prompt new drug prescription(s), known as prescribing cascades. We aimed to identify potential ARB-induced prescribing cascades using high-throughput sequence symmetry analysis.

**Methods:** Using claims data from a national sample of Medicare beneficiaries (2011–2020), we identified new ARB users aged ≥66 years with continuous enrollment ≥360 days before and ≥180 days after ARB initiation. We screened for initiation of 446 other (non-antihypertensive) ‘marker’ drug classes within ±90 days of ARB initiation, generating sequence ratios (SRs) reflecting proportions of ARB users starting the marker class after versus before ARB initiation. Adjusted SRs (aSRs) accounted for prescribing trends over time, and for significant aSRs, we calculated the naturalistic number needed to harm (NNTH); significant signals were reviewed by clinical experts for plausibility.

**Results:** We identified 320,663 ARB initiators (mean ± SD age 76.0 ± 7.2 years; 62.5% female; 91.5% with hypertension). Of the 446 marker classes evaluated, 17 signals were significant, and three (18%) were classified as potential prescribing cascades after clinical review. The strongest signals ranked by the lowest NNTH included benzodiazepine derivatives (NNTH 2130, 95% CI 1437–4525), adrenergics in combination with anticholinergics, including triple combinations with corticosteroids (NNTH 2656, 95% CI 1585–10074), and other antianemic preparations (NNTH 9416, 95% CI 6606–23784). The strongest signals ranked by highest aSR included other antianemic preparations (aSR 1.7, 95% CI 1.19–2.41), benzodiazepine derivatives (aSR 1.18, 95% CI 1.08–1.3), and adrenergics in combination with anticholinergics, including triple combinations with corticosteroids (aSR 1.12, 95% CI 1.03–1.22).

**Conclusion:** The identified prescribing cascade signals reflected known and possibly under-recognized ARB adverse events in this Medicare cohort. These hypothesis-generating findings require further investigation to determine the extent and impact of these prescribing cascades on patient outcomes.

## INTRODUCTION

Angiotensin II receptor blockers (ARBs) have been developed to selectively inhibit the renin-angiotensin-aldosterone system (RAAS) by blocking AT1 receptors, the final step in the RAAS cascade. Although ARBs share clinical indications with angiotensin-converting enzyme inhibitors (ACEI), they differ pharmacologically. Angiotensin-converting enzyme inhibitors reduce the production of angiotensin II, thereby indirectly affecting binding to both type 1 (AT1) and type 2 (AT2) receptors.(1–7) In contrast, ARBs selectively block AT1 receptors without altering the overall production of angiotensin II and, unlike ACEIs, do not increase bradykinin levels.(8–10)

Studies have shown that ARBs and ACEIs have similar effectiveness in lowering blood pressure, with ARBs providing an added benefit of favorable safety profiles, including lower risk of cough, angioedema, gastrointestinal bleeding, and acute pancreatitis, and potential benefits in preventing cognitive decline.(11–18) Despite differences in safety profiles, hypertension guidelines recommend ACEIs and ARBs as primary treatment options for hypertension in adults.(19–21) However, ARBs are preferred over ACEI for patients with chronic pulmonary diseases and coughs. Like ACEIs, ARBs are also used in the treatment of heart failure and chronic kidney disease.(22–25) As of 2019, ARBs accounted for 17% of the antihypertensives dispensed in the United States, with their use as initial agents in the treatment of incident hypertension has increased significantly over the last decade.(26)

Although ARBs have been available since 1995, most data on their adverse drug event (ADE) profile come from randomized controlled trials (RCTs), which indicate that ARBs are generally well tolerated.(27–30) However, reported adverse events include angioedema, cough, headache, dizziness, fatigue, and hyperkalemia.(31–35) If undetected or unknown, adverse events caused by ARBs could prompt the prescription of a new medication to manage ARB-related adverse events, a phenomenon known as a prescribing cascade. (36) Given their widespread use in diverse patient populations, many with multiple comorbidities, the potential impact of ARB-related prescribing cascades could be substantial.

To address this knowledge gap, high-throughput screening of prescribing cascades in a diverse, real-world population of older adults, who are often underrepresented in RCTs, is needed. Such approach would capture not only well-known ARB-related adverse events but also identify new ones. Identifying these prescribing cascades is essential for informing deprescribing initiatives and mitigating risks associated with polypharmacy. Therefore, we conducted high-throughput signal detection using sequence symmetry analysis (SSA) to identify potential ARB-related prescribing cascades among Medicare beneficiaries.

## METHODS

### Data sources

We used claims data from a 5% national sample of Medicare beneficiaries with fee-for-service (FFS) coverage from to 2011-2015 plus 1 million FFS beneficiaries in Florida and a 15% national sample of Medicare FFS beneficiaries plus all FFS beneficiaries in Florida from to 2016-2020. Medicare is a US federal insurance program for adults aged ≥65 years and others with qualifying conditions. Medicare FFS includes Part A (hospital), Part B (medical), and Part D (prescription drug) services, which capture inpatient and outpatient services, pharmacy claims, and beneficiary characteristics. The study was considered exempt by the University of Florida’s Institutional Review Board. We used Strengthening the Reporting of Observational Studies in Epidemiology (STROBE) reporting guidelines to ensure appropriate reporting.(37)

### Design

High-throughput SSA was used to identify potential ARB-related prescribing cascades. (38–40) PSSA is a hypothesis-free pharmacovigilance approach that employs a case-only study design to assess the temporality of an “index” drug initiation (i.e., ARB) relative to initiation of all other “marker” class (i.e., all other medication classes), hereafter referred to as ARB-marker class dyads.(41, 42) Included patients were ARB initiators who had their first ARB fill between 2011 and 2019, were aged ≥66 years at ARB initiation, and had ≥360 days of continuous insurance coverage before (parts A, B, and D) and ≥180 days after ARB initiation. We required continuous insurance coverage to ensure genuine new use of ARB and to capture all marker class use during the 180 days prior to and after ARB initiation.

Anatomical Therapeutic Chemical (ATC) codes were used to hierarchically group marker drugs into medication classes for high-throughput screening. The ATC classification system is maintained by the World Health Organization and classifies medications into five levels: Level 1 indicates the broad anatomical group (n = 14 total), Level 4 indicates the chemical subgroup/drug class, and Level 5 indicates the specific drug/chemical substance (n ≈ 5000 total) (Supplemental Table S1).(43) Among the ARB initiators, we identified the first claim for any marker class within a given ATC Level 4 subgroup. If an individual filled out multiple different medications (unique ATC Level 5) within a given ATC Level 4 subgroup during the study period, we only included the first fill within that ATC Level 4 subgroup. We required the marker class initiation to occur within ±90 days of ARB initiation for the primary analysis to focus on acute onset adverse events, and we excluded all patients initiated on other classes of antihypertensive drugs on the same day as ARB initiation. (44) In sensitivity analyses, we extended the marker drug initiation time window to ±180 days to allow the identification of adverse events (and subsequent prescribing cascades) with longer induction periods.

For each ATC Level 4 subgroup, all included patients in the ARB marker class dyad were evaluated using the PSSA methodology. These analyses were completed iteratively until all ATC Level 4 subgroups were evaluated, excluding ATC Level 4 subgroups representing antihypertensive classes. Baseline characteristics (age, sex, calendar year of ARB initiation, Charlson Comorbidity Index, specific ARB medication, and other comorbidities) of ARB initiators were measured at the time of ARB initiation or 360 days prior to ARB initiation.(45, 46)

### Analyses

For each unique ARB marker class dyad, we determined the crude sequence ratio (cSR) as the number of patients who initiated the marker class after ARB initiation divided by the number of patients who initiated the marker class before the ARB. Excess initiation of a marker class after ARB, relative to before ARB, results in a cSR >1 and may indicate a prescribing cascade. To adjust for background prescribing trends, we estimated an adjusted sequence ratio (aSR) with 95% confidence intervals (CI) by dividing the cSR by the null-effect ratio for each ARB-marker class dyad.(47) All aSRs with a lower CI limit >1 were considered significant signals for further exploration under the assumption that no within-person time-varying confounding exists.

Each ARB-marker class dyad was represented graphically by plotting the distribution of the timing of marker class initiation (in 10-day intervals) relative to ARB initiation for each exposure window (±90 and ±180 days) surrounding ARB initiation. In addition, for each ARB-marker class dyad considered statistically significant, we estimated the excess risk among exposed individuals and the corresponding naturalistic number needed to harm (NNTH) within 1 year. Excess risk was calculated as the difference between the number of patients initiated on the marker class after ARB initiation and the number of patients initiated on the same marker class before ARB initiation, divided by the total number of ARB initiators, standardized to a rate per 1000 person-years accounting for a 90-day exposure window in the primary analysis. The NNTH was calculated as the inverse of the excess risk among the exposed, that is, the number of patients who needed to be treated with an ARB for one additional patient to experience one prescribing cascade within that marker class.

### Classification of signals

All statistically significant signals (aSRs with 95% CIs >1) in the primary analysis were manually reviewed through a systematic process to differentiate potential prescribing cascades from false positive signals. False-positive signals could be attributable to detection bias (i.e., a new condition identified with a corresponding new medication initiated during routine monitoring of the ARB), disease progression (i.e., new medications initiated to treat the progression of underlying cardiovascular disease), therapeutic escalation (i.e., escalation of therapy [2nd or 3rd line treatments] in conditions related to ARB indication), or reverse causation (i.e., decreased ARB initiation following the initiation of marker class [e.g., reduced ARB treatment following late-stage cancer treatment]). Signals not indicative of potential prescribing cascades were classified as “Other,” as these false-positive signals may be due to multiple biases.

Manual review was conducted in two stages. First, pharmacy trainees (n=3) were trained by study investigators (SMS & KMS) in the use of primary (e.g., PubMed/MEDLINE searches), secondary (e.g., drug monographs, package inserts), and tertiary (e.g., drug information databases) drug information sources for assessing potential prescribing cascades and their underlying mechanisms to support signal classification. Each significant signal was assigned to the three pharmacy trainees, who independently reviewed the signals and assigned an initial classification, as described above, along with supporting literature. Second, two pharmacists with clinical expertise in medication and patient safety then independently classified each signal using material developed by pharmacy trainees and *ad-hoc* literature evaluations when needed. In cases of disagreement between the clinically trained pharmacists, consensus was reached by the same clinical pharmacists together with senior study investigators (SMS, EJM).

All analyses were conducted using SAS statistical software (version 9.4; SAS Institute, Cary, NC, USA) and visualized with R (2023.09.1) and Tableau (2023.06.2+561).

## RESULTS

We identified 320,663 ARB initiators; their baseline characteristics are summarized in the Table. Briefly, 62.5% of the patients were women, and the mean age (± SD) was 76 ± 7.2 years. The majority (78.8%) were Non-Hispanic White, 7.3% were black, and 3.4% were Asian/Pacific Islanders. Most patients (75.7%) had a Charlson Comorbidity Index score ≥5. Losartan was the most commonly initiated ARB (77.6%) followed by valsartan (15.2%).

All high-throughput screening results are interactively displayed at: https://public.tableau.com/app/profile/cvmedlab/viz/ARBs_newdata/TableofContentsFlowchart2, with significant aSRs presented in Supplemental Table S2, non-significant signals in Supplemental Table S3, and the result of all sensitivity analyses in Supplemental Table S4. For the primary analysis, among 446 marker drug classes analyzed, we found 17 statistically significant signals (Figure 1 & 2). Among these 17 significant signals, only three were classified as potential prescribing cascades following a clinical review: other antianemic preparations (B03XA), benzodiazepine derivatives (N03AE), and adrenergics in combination with anticholinergics, including triple combinations with corticosteroids (R03AL) (Figure 1). Figure 2 summarizes the statistically significant signals ranked by the lowest NNTH.

**Figure 1.**
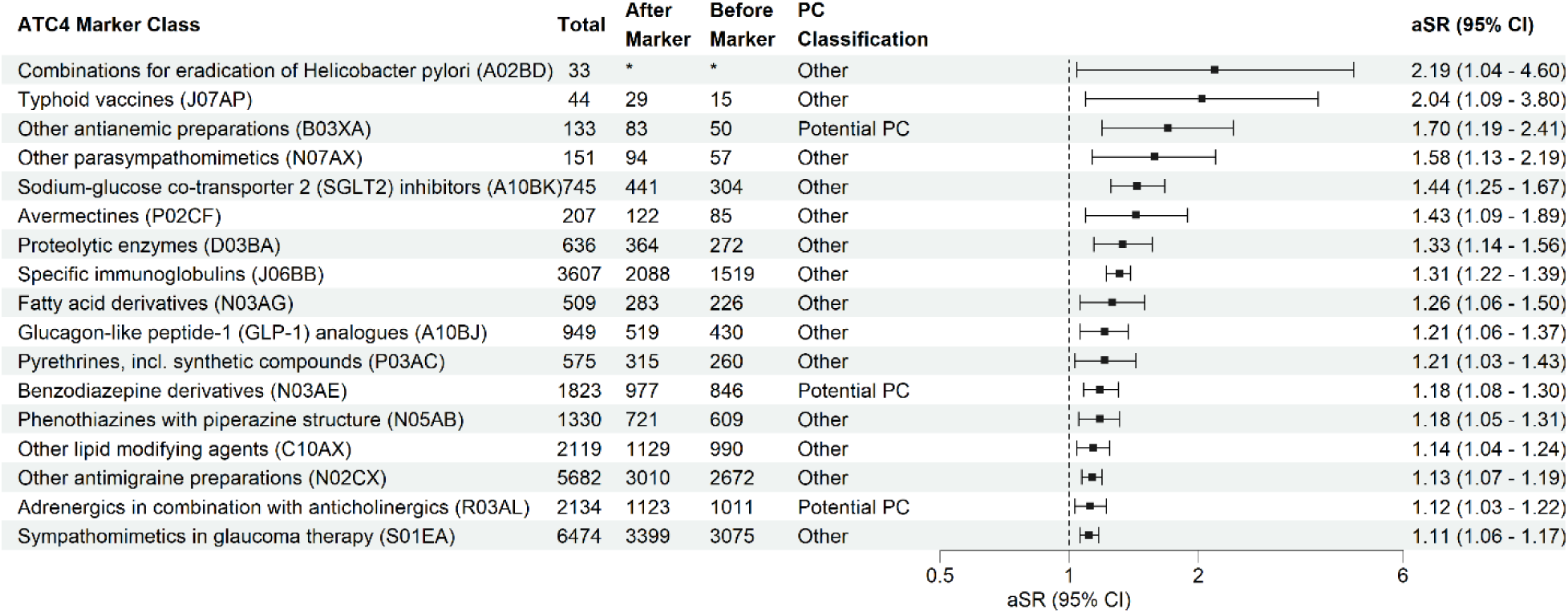
Significant signals from sequence symmetry analyses of ARB-marker class dyads by adjusted sequence ratio. ATC4, Anatomical Therapeutic Chemical - Level 4, NNTH, Naturalistic Number Needed to Harm (within 1 year); PC, Prescribing Cascade CI, Confidence Interval.

**Figure 2.**
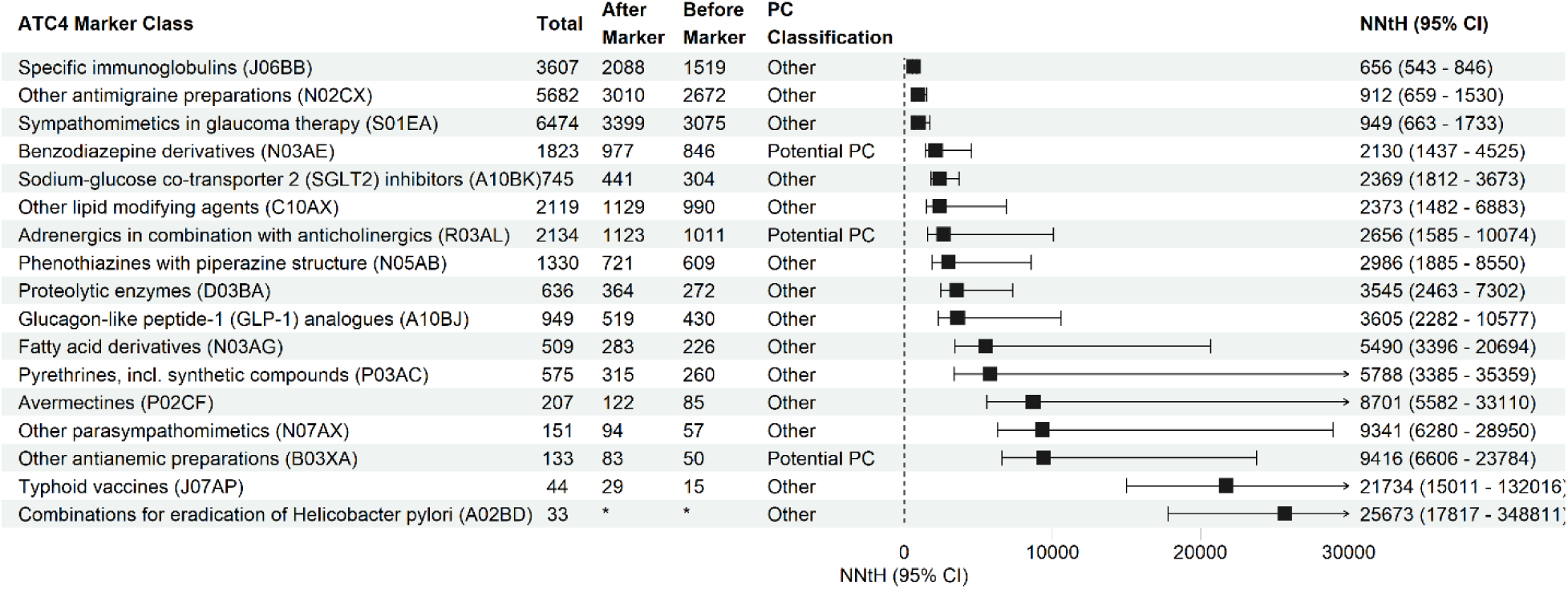
Significant signals from sequence symmetry analyses of ARB-marker class dyads by naturalistic number need to harm. ATC4, Anatomical Therapeutic Chemical - Level 4, NNtH, naturalistic number needed to harm (within 1 year); PC, Prescribing Cascade CI, confidence interval.

Figure 3 summarizes the statistically significant ARB inhibitor-marker class dyads by NNTH and aSR, and highlights those that were classified as potential prescribing cascades. All potential prescribing cascades had an aSR <2 and NNTH<10,000.

**Figure 3.**
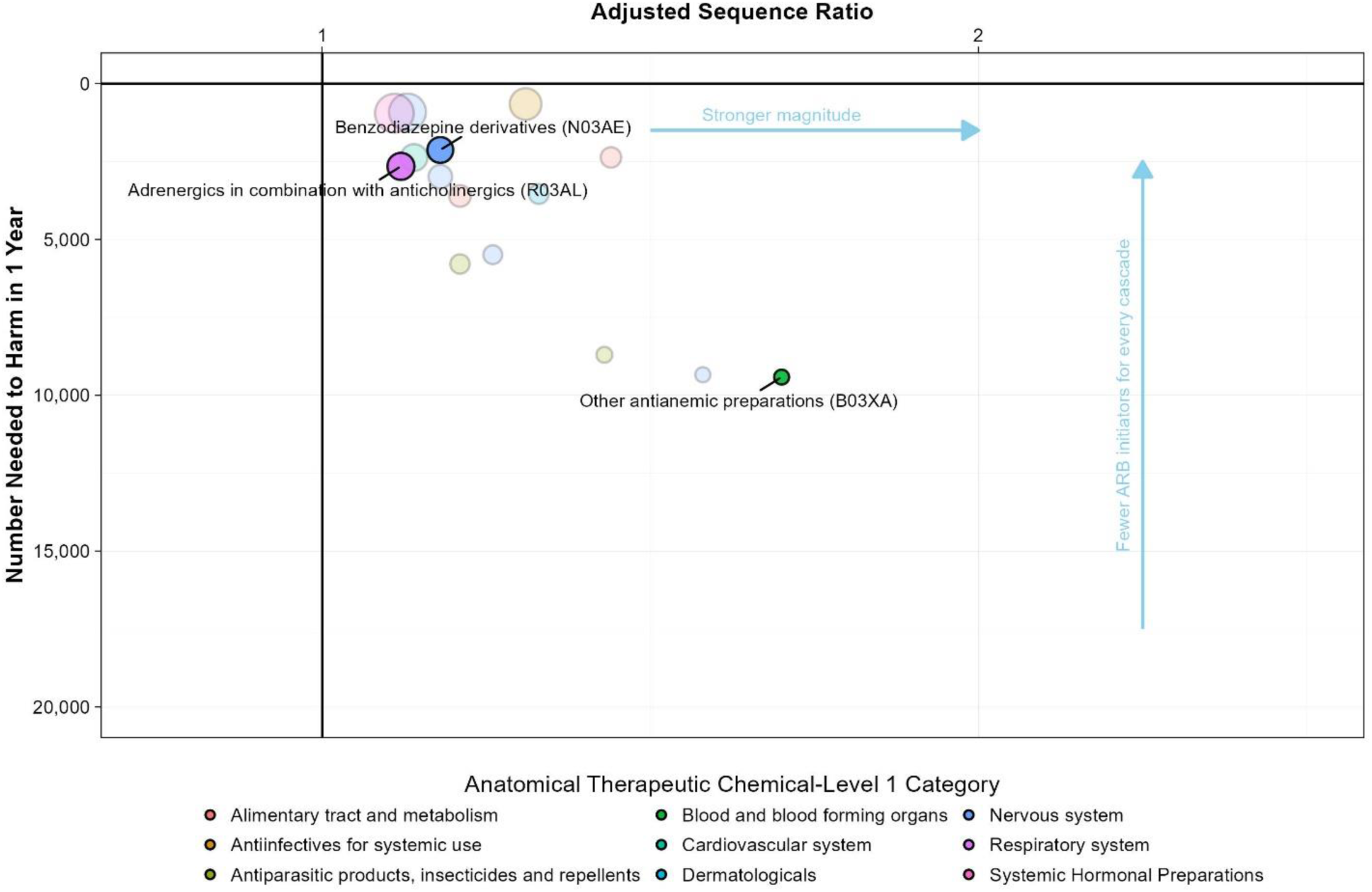
Significant ARB inhibitor-marker class dyad signals by adjusted sequence ratio and naturalistic number needed to harm. Dyads are grouped (color-coded) at the Anatomical Therapeutic Chemical Level 1 category. The faded dots are dyads classified as ‘other’ while the colored dots are dyads classified as potential prescribing cascades weighted by the total number of prescriptions. All results from the high-throughput screening are displayed interactively at https://public.tableau.com/app/profile/cvmedlab/viz/ARBs_newdata/TableofContentsFlowchart2

## DISCUSSION

Using population-based high-throughput prescription sequence symmetry analysis (SSA), we identified medication classes that were prescribed more frequently after ARB initiation than before. Among the 446 marker classes evaluated, we identified 17 significant signals, of which three were considered potential ARB prescribing cascades after clinical review. Signals were ranked based on (1) aSR, as a measure of the magnitude of the signal, and (2) NNTH or excess risk to the exposed, as a measure of the impact within the population. When determining which prescribing cascades have a significant clinical impact on most patients, NNTH or excess risk among exposed patients is straightforward and useful for clinical decision-making and has been used in prior high-throughput SSA screening evaluation.(41)

Ranking the signals using NNTH, the strongest signal classified as a potential prescribing cascade was benzodiazepine derivatives (N03AE). An association between RAS inhibitors and depression or anxiety has been reported in several studies (48–50). Recent studies have highlighted the link between the renin-angiotensin system and mood disorders, emphasizing the role of Angiotensin II-induced Nicotinamide Adenine Dinucleotide Phosphate (NADPH) oxidase-derived oxidative stress in the central nervous system as a potential mechanism underlying the pathogenesis of these conditions.(51) Mamdani et al. reported a nested case-control study using real-world data that found that patients receiving ARBs had a greater risk of suicide than those receiving ACEIs.(49) Our findings contribute to the growing body of evidence and highlight the need for further rigorous study designs, including randomized clinical trials, to confirm these observations and better inform clinical prescription practices.

Although less prevalent than in ACEIs, ARBs can cause life-threatening angioedema. (52, 53) ARB-induced angioedema could explain the excess prescription of salbutamol, formoterol and fluticasone (adrenergics in combination with anticholinergics [R03AL]) for treating bronchospasm and other upper airway manifestation.(54–56) While the incidence of angioedema is generally lower with ARBs than with ACEIs, there is a need for ongoing post-marketing surveillance to better define the true incidence of angioedema associated with ARBs and its potential for downstream prescribing cascades.

Angiotensin II receptor blockers, particularly losartan, may decrease erythropoietin (57–66). The adverse effects are heightened in the presence of chronic kidney disease due to the accumulation of N-acetyl-seryl-aspartyl-lysyl-proline.(57) The resulting suppression of erythropoiesis may trigger a prescribing cascade involving antianemic preparations (B03XA) such as erythropoietin and darbepoetin alfa. Monitoring hematologic parameters in patients on ARBs, particularly those with chronic kidney disease or undergoing kidney transplant, is key to mitigating risks.

In one of our studies using the same patient population, we identified 42 potential prescribing cascades among ACEI initiators compared to only three among ARB initiators.(67) Research has consistently demonstrated that ARBs have a more favorable safety profile, with patients experiencing lower risks of angioedema and cough among other side effects.(11–18) The observed differences in prescribing cascades between ACEIs and ARBs could be due to the mechanism of action and clinical practice or a combination of both. Mechanistically, ACEIs inhibit ACE and reduce angiotensin II levels, but increase bradykinin, which is associated with adverse effects such as dry cough and angioedema.(1–7, 28, 68–84) Such adverse events lead to prescribing cascades involving antitussives, bronchodilators, or corticosteroids.(67, 85–88) In contrast, ARBs block angiotensin II type 1 receptors without altering bradykinin levels, resulting in fewer side effects and, consequently, fewer prescribing cascades. (8–10) Clinical practice patterns also contribute to this difference. Angiotensin-converting enzyme inhibitors have been on the market for a longer time, leading to greater familiarity among physicians, who may prescribe them more frequently and address their adverse effects with additional medications, whereas the milder adverse events associated with ARBs often require fewer interventions. (89, 90) In addition, ARBs are often prescribed to patients who have previously experienced ACEI-related adverse events, leading to closer monitoring and stricter health surveillance. (91–93) For older adults at high risk of polypharmacy, an additional advantage of ARBs is their potential to minimize the risk of prescribing cascade and its downstream consequences. Our findings add to the growing body of evidence supporting ARBs as a preferred first-line option over ACEIs for hypertension management owing to a favorable safety profile.(16, 94)

After reviewing the 17 identified signals, we classified 14 (82%) as unlikely to be a prescription cascade. Several factors contributed to this classification, including the pharmacological profile of the medication, the expected timeframe for an adverse event to occur after the index drug was used, the possibility of reverse causation, and whether the signal was perceived to be more likely due to disease progression rather than managing an adverse drug event. For example, signals observed for various diabetes medications, such as sodium-glucose co-transporter 2 (SGLT2) inhibitors (A10BK) and glucagon-like peptide-1 (GLP-1) analogs (A10BJ), were thought to be attributed more to disease progression or confounding by indication. Hypertension is an independent risk factor for diabetes(95–97) and individuals with or at risk for diabetes may be selectively prescribed RAS inhibitors to reduce the risk of macro- and microvascular events. Additionally, we identified signals such as specific immunoglobulins (e.g., varicella/zoster immunoglobulin varicella/zoster immunoglobulin) and tetanus vaccines (J07AM), which were perceived to be likely due to detection bias. For example, these observed signals could be due to enhanced health surveillance that occurs when patients begin new medications and are hence more frequently monitored within the health system.

Key strengths of the study include being the first high-throughput SSA screening for prescribing cascades associated with ARBs conducted in the US, and focusing on one of the most prescribed drug classes using a nationally representative sample of the older population. We classified all significant signals and used an approach that allowed the assessment of signals affecting relatively few patients. In addition, we publicly report our aggregate findings, allowing for transparency.

Our study also had important limitations. Signal classification was based on data and clinical knowledge and may be subject to misclassification, though multiple evaluators and consensus review were used. The findings may be overestimated due to within-person time-varying bias, such as disease progression or a new diagnosis, which we attempted to minimize by restricting marker initiation to a 90-day exposure around ARB initiation. The cohort consisted of Medicare beneficiaries, potentially limiting generalizability to younger populations or those with different health insurance coverage. Importantly, these findings are hypothesis generating, and additional confirmation in a traditional cohort study is needed. Future studies should incorporate negative controls to reduce within-patient time-varying confounding factors such as hypertension progression or age-related changes.(98–100) Finally, we did not adjust for multiple testing, which could increase the risk of spurious associations; however, previous studies suggest such corrections may not always be optimal.(40, 41, 101, 102)

All potential prescribing cascades identified in this study require validation in well-designed cohort studies, with priority given to medication classes classified as potentially inappropriate for older adults by the Beers Criteria, those with low NNTH values, and those frequently prescribed to large patient populations.(103) After validation, studies should assess the risks and benefits of the prescribing cascades to determine their clinical appropriateness and identify potentially problematic prescribing cascades.(104–106) Further evaluations should examine clinical relevance, identify predictors and characterize high-risk subpopulations. Future research could also investigate drug classes prescribed less frequently following ARB initiation to identify opportunities for drug repurposing.

## CONCLUSION

Using high-throughput SSA screening in a population of Medicare beneficiaries, we identified previously known prescribing cascades, new potential prescribing cascades based on known ARB adverse events, and new potential prescribing cascades based on previously unknown adverse events. While this approach to identifying prescribing cascades should be considered hypothesis generating, our findings could initiate discussions in clinical settings to ensure that the benefits of ARB are optimized while minimizing the risks of potentially harmful prescribing practices.

## Abbreviations Definition List

ACEI: Angiotensin-Converting Enzyme Inhibitor
ADE: Adverse Drug Event
ARB: Angiotensin II Receptor Blocker
aSR: Adjusted Sequence Ratio
CCI: Charlson Comorbidity Index
CI: Confidence Interval
FFS: Fee-for-Service
NNTH: Number Needed to Harm
PC: Prescribing Cascade
RAAS: Renin-Angiotensin-Aldosterone System
RCT: Randomized Controlled Trial
SSA: Sequence Symmetry Analysis
STROBE: Strengthening the Reporting of Observational Studies in Epidemiology
WHO ATC: World Health Organization Anatomical Therapeutic Chemical Classification System

## Ethics Statement

The study was approved as Exempt by the University of Florida Institutional Review Board.

## Data Availability Statement

The study was conducted using CMS Medicare Fee-for-Service claims databases pursuant to a data-use agreement between the University of Florida and CMS, which prevents the sharing of data entrusted to the University of Florida. However, qualified researchers can obtain such data directly from the CMS. The SAS code used in this study is available from the Corresponding Author on request.

## AUTHOR CONTRIBUTIONS

Asinamai M Ndai contributed to study design and drafted the manuscript. Smith, Morris, and Vouri conceived and designed the study and secured funding. Earl J. Morris, Asinamai M. Ndai, Shailina Keshwani, Scott M. Vouri, Kayla Smith, and Steven M. Smith were responsible for statistical analysis and visualization of the data. All authors made substantial contributions to the interpretation of the data and results, reviewed and provided critical revisions, and approved the final manuscript.

---

**Table.**
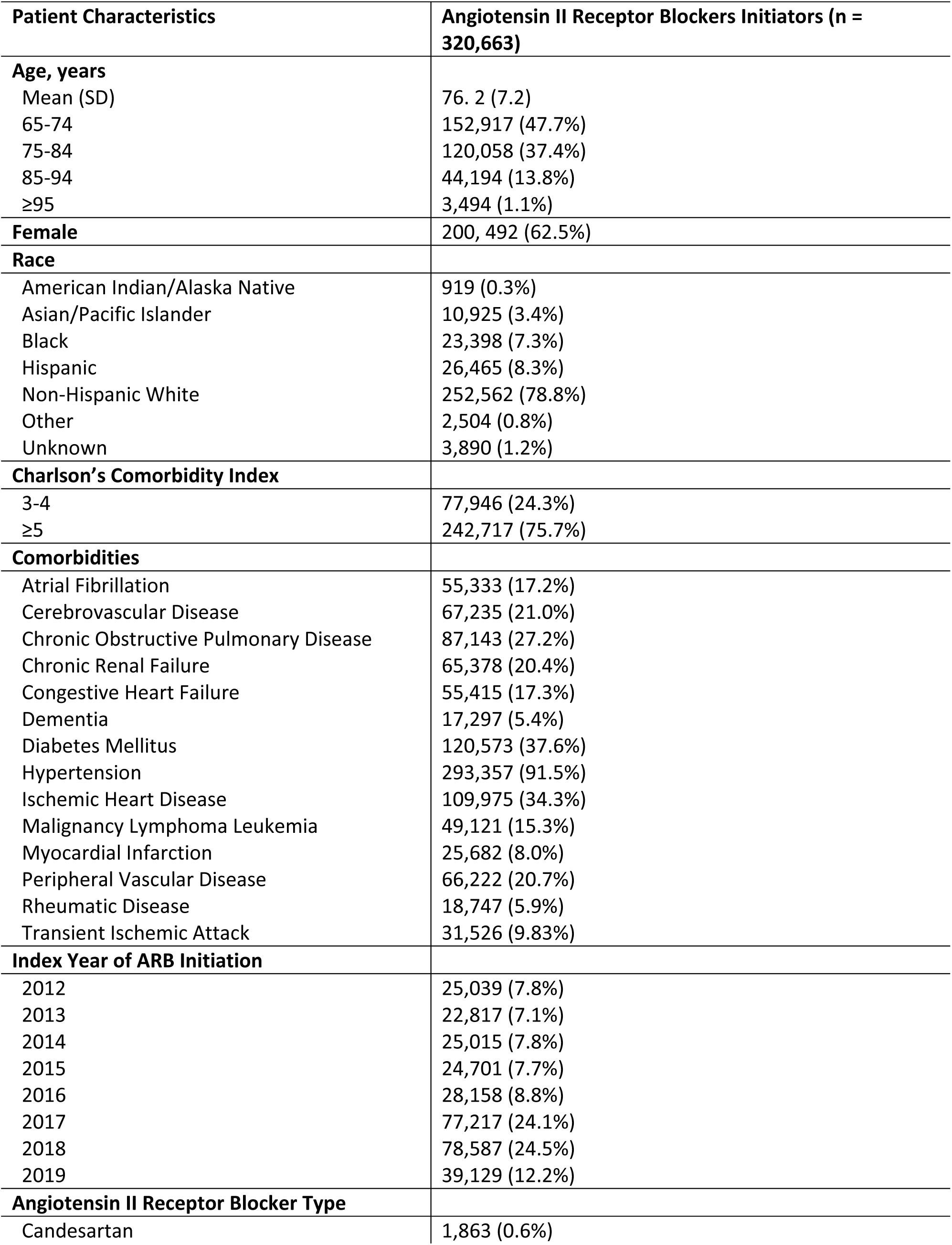
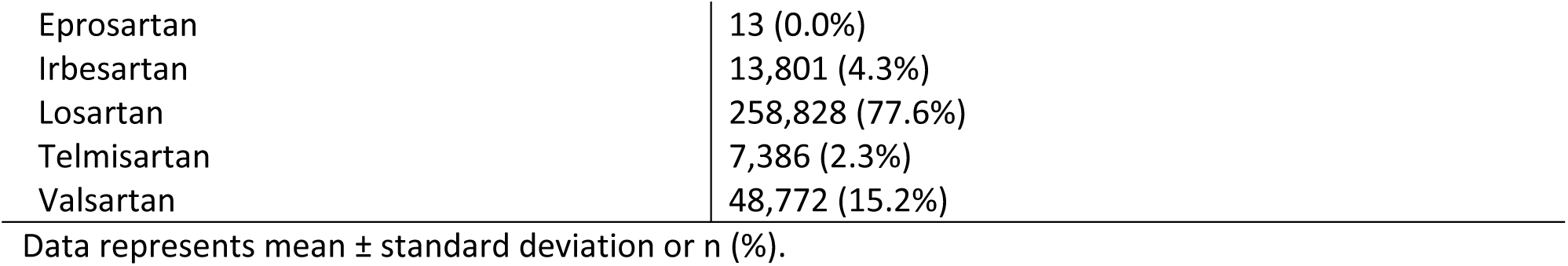
Baseline characteristics of ARB initiators included in the cohort.

| **Patient Characteristics** | **Angiotensin II Receptor Blockers Initiators (n = 320,663)** |
| --- | --- |
| **Age, years** |  |
| Mean (SD) | 76.2 (7.2) |
| 65-74 | 152,917 (47.7%) |
| 75-84 | 120,058 (37.4%) |
| 85-94 | 44,194 (13.8%) |
| ≥95 | 3,494 (1.1%) |
| **Female** | 200,492 (62.5%) |
| **Race** |  |
| American Indian/Alaska Native | 919 (0.3%) |
| Asian/Pacific Islander | 10,925 (3.4%) |
| Black | 23,398 (7.3%) |
| Hispanic | 26,465 (8.3%) |
| Non-Hispanic White | 252,562 (78.8%) |
| Other | 2,504 (0.8%) |
| Unknown | 3,890 (1.2%) |
| **Charlson's Comorbidity Index** |  |
| 3-4 | 77,946 (24.3%) |
| ≥5 | 242,717 (75.7%) |
| **Comorbidities** |  |
| Atrial Fibrillation | 55,333 (17.2%) |
| Cerebrovascular Disease | 67,235 (21.0%) |
| Chronic Obstructive Pulmonary Disease | 87,143 (27.2%) |
| Chronic Renal Failure | 65,378 (20.4%) |
| Congestive Heart Failure | 55,415 (17.3%) |
| Dementia | 17,297 (5.4%) |
| Diabetes Mellitus | 120,573 (37.6%) |
| Hypertension | 293,357 (91.5%) |
| Ischemic Heart Disease | 109,975 (34.3%) |
| Malignancy Lymphoma Leukemia | 49,121 (15.3%) |
| Myocardial Infarction | 25,682 (8.0%) |
| Peripheral Vascular Disease | 66,222 (20.7%) |
| Rheumatic Disease | 18,747 (5.9%) |
| Transient Ischemic Attack | 31,526 (9.83%) |
| **Index Year of ARB Initiation** |  |
| 2012 | 25,039 (7.8%) |
| 2013 | 22,817 (7.1%) |
| 2014 | 25,015 (7.8%) |
| 2015 | 24,701 (7.7%) |
| 2016 | 28,158 (8.8%) |
| 2017 | 77,217 (24.1%) |
| 2018 | 78,587 (24.5%) |
| 2019 | 39,129 (12.2%) |
| **Angiotensin II Receptor Blocker Type** |  |
| Candesartan | 1,863 (0.6%) |
| Eprosartan | 13 (0.0%) |
| Irbesartan | 13,801 (4.3%) |
| Losartan | 258,828 (77.6%) |
| Telmisartan | 7,386 (2.3%) |
| Valsartan | 48,772 (15.2%) |

Data represents mean ± standard deviation or n (%).

